# The Harvard-Emory ECG Database

**DOI:** 10.1101/2024.09.27.24314503

**Authors:** Zuzana Koscova, Qiao Li, Chad Robichaux, Valdery Moura Junior, Manohar Ghanta, Aditya Gupta, Jonathan Rosand, Aaron Aguirre, Shenda Hong, David E. Albert, Joel Xue, Aarya Parekh, Reza Sameni, Matthew A. Reyna, M. Brandon Westover, Gari D. Cliford

## Abstract

The Harvard-Emory ECG Database (HEEDB) is a large collection of 12-lead electrocardiogram (ECG) recordings, developed through a collaboration between Harvard University and Emory University. The database consists of 10,608,417 unique ECG recordings from 1,818,247 patients from Massachusetts General Hospital (MGH) and 1,452,964 recordings from 552,481 patients from Emory University Hospital (EUH) collected in clinical settings as part of routine patient care since the early 1990s. Continuously updated with new data, the recordings consist of 10-second, 12-lead ECGs sampled at 250 and 500 Hz, and stored in WFDB format. Future updates will include demographic information such as age, sex, race, and ethnicity. Additionally, 12SL annotations–encompassing ECG diagnoses, morphology, and rhythms–are available for 10,471,531 recordings from MGH and 1,268,277 recordings from EUH. Shortly, ICD-9/10 codes, CPT codes, and medication history will also be made publicly available.

## Background & Summary

The Harvard-Emory ECG database (HEEDB) is a large collection of 12-lead (I, II, III, aVR, aVL, aVF, V1, V2, V3, V4, V5, V6) electrocardiography (ECG) recordings, prepared through a collaboration between Harvard University and Emory University investigators. HEEDB comprises clinical ECGs obtained during routine care over several decades (from the 1990s to present), alongside associated covariates, including patient demographics, medications, diagnoses, and clinical outcomes which will be added in the future. These data were collected at Massachusetts General Hospital (MGH, Boston, MA, US) and Emory University Hospital (EUH, Atlanta, GA, US) and represent one of the largest ECG datasets available. Large-scale datasets with extended follow-up, such as this one, are uncommon, making this resource particularly valuable for conducting population-wide analyses of cardiac rhythms, their disturbances, and associated outcomes.

The HEEDB supports numerous applications, ranging from identifying early markers of cardiovascular disease to validating detection and prediction algorithms for clinical use. A major focus is the identification of ECG-derived biomarkers that may predict arrhythmias, sudden cardiac death, and other critical conditions. The inclusion of temporal data, with multiple ECG recordings for individual subjects over time, provides an opportunity to study the progression of cardiac abnormalities and their clinical implications. The scale and longitudinal nature of this database offer significant potential to advance both clinical research and computational cardiology, enabling new insights into cardiovascular disease.

## Methods

The HEEDB contains 12-lead ECG recordings of 10 seconds duration sampled at 250 and 500 Hz. At the time of initial publication, the database includes 10,608,417 unique ECG recordings (10,771,552 ECGs total) from 1,818,247 patients from MGH. New ECGs will be added to the database periodically, including 1,452,964 recordings from 552,481 subjects recorded at EUH. All ECGs were collected in the course of routine clinical care using the MUSE ECG system. The MUSE system facilitates the acquisition, storage, and review of ECG data, providing consistent recordings across the dataset. The raw ECG waveforms, along with associated metadata, were stored in the XML format, which allows for both structured data representation and interoperability with other systems. ECG waveforms were further de-identified following the Safe Harbor method and converted to the WFDB (Waveform Database)^1,2^and MATLAB (V4) compatible format.

## Data Records

Each ECG recording includes one waveform data file (.mat) and one header file (.hea). The waveform data file can be read by WFDB library functions, applications, and Toolbox, or be loaded to MATLAB directly. Most waveform files are synchronized 12-lead ECG signals recorded at 250 Hz (59% from MGH) and 500 Hz (41% from MGH, 100% from EUH) for a duration of 10 s with analog-to-digital converter (ADC) gain 1,000, stored in mV with byte offset of 24. The header file specifies the names of the associated waveform files and their attributes such as sampling rate and units, including the channel names of the signal. It contains line-oriented and field-oriented ASCII text and can be read by the WFDB library or generic text editors. The data file contains 12-lead information encoded in 16 bits. The ECG database exhibits several common abnormalities that occur in real ECG recordings, such as missing leads, incomplete time lengths, and various types of noise affecting the ECG recordings.

### Metadata

The dataset includes general metadata for each subject, including a de-identified unique subject identifier, sex, age, race, ethnicity, and a shifted acquisition date. All the dates are shifted between *±* 90 days from the date of acquisition and the shift is different for every subject/recording. Demographic characteristics for both data sources are summarized in Table 1. To ensure confidentiality, individuals aged over 89 years at the time of data acquisition are uniformly recorded as 90 years old. It is important to note that age data is missing for 15,466 recordings from MGH and 37,481 recordings from EUH due to discrepancies in documenting either the acquisition date or birth date. The age distribution for both data sources is displayed in the histogram in Figure 1.

**Table 1.** Available demographic information for the ECG records in the HEEDB dataset, including age, gender, race, ethnicity, and education level.

| HEEDB | MGH |  | Emory |  |
| --- | --- | --- | --- | --- |
|  | Subjects<br>Total (n=1,818,247) | Recordings<br>Total (n=10,608,417) | Subjects<br>Total (n=552,481) | Recordings<br>Total (n=1,452,964) |
| **Age** |  |  |  |  |
| Age, years (mean (std)) | - | 60.61 (18.3) | - | 61.4 (16.1) |
| Unavailable | - | 15,466 (0.2%) | - | 37,481 (2.6%) |
| - [0 – 2) | - | 33,907 (0.3%) | - | 500 (0.03%) |
| - [2 – 10) | - | 64,837 (0.6%) | - | 215 (0.01%) |
| - [10 – 18) | - | 114,391 (1.1%) | - | 3,900 (0.3%) |
| - [18 – 40) | - | 1,309,892 (12.4%) | - | 160,398 (11.0%) |
| - [40 – 60) | - | 3,027,640 (28.5%) | - | 446,091 (30.7%) |
| - [60 – 89) | - | 5,736,417 (54.1%) | - | 776,837 (53.5%) |
| - >89 | - | 305,867 (2.9%) | - | 27,542 (1.9%) |
| **Sex** |  |  |  |  |
| Female | 925,510 (51.1%) | - | 284,979 (51.9%) | - |
| Male | 886,155 (48.7%) | - | 266,955 (48.3%) | - |
| Unavailable | 3,582 (0.2%) | - | 547 (0.1%) | - |
| **Race** |  |  |  |  |
| White | 1,291,671 (71.0%) | - | 287,707 (52.1%) | - |
| Unavailable | 347,943 (19.1%) | - | 65,774 (11.9%) | - |
| Black or African American | 120,207 (6.6%) | - | 183,456 (33.2 %) | - |
| Asian | 49,748 (2.7%) | - | 12,366 (2.2%) | - |
| Other | 8,698 (0.4%) | - | 3,178 (0.6%) | - |
| **Ethnicity** |  |  |  |  |
| Non-Hispanic | 844,564 (46.5%) | - | - | - |
| Unavailable | 848,415 (46.7%) | - | - | - |
| Hispanic | 125,268 (6.9%) | - | - | - |
| **Education** |  |  |  |  |
| Unavailable | 861,933 (47.4%) | - | - | - |
| Did not complete high school | 97,953 (5.4%) | - | - | - |
| High school or equivalent | 334,142 (18.4%) | - | - | - |
| Some college | 82,648 (4.6%) | - | - | - |
| College and/or advanced degree | 504,120 (27.7%) | - | - | - |

**Figure 1.**
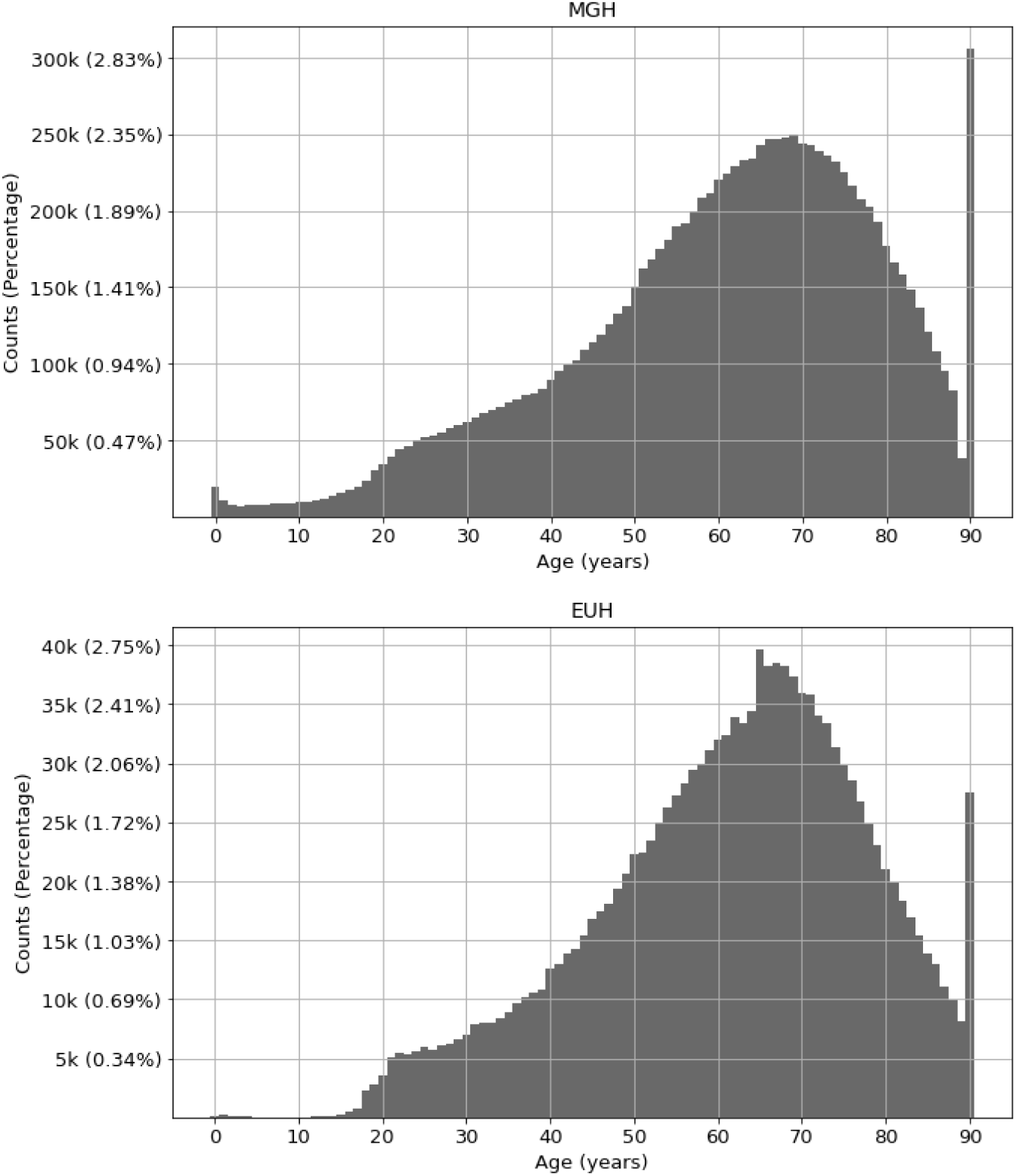
Histogram depicting the age distribution within the HEEDB dataset for Massachusetts General Hospital (MGH) recordings (top figure) and Emory University Hospital (EUH) recordings (bottom figure).

### ECG Annotations

The annotations in the dataset are generated using the Marquette 12SL ECG Analysis Program (GE Healthcare) version 4^3^. These annotations consist of textual reports that describe the morphology, rhythm, and diagnostic information of the ECGs. Each annotation includes statement numbers that correspond to human-readable diagnoses. The frequency of statements with the highest occurrence is summarized in Table 2 for MGH and Table 3 for EUH.

**Table 2.** This table lists the 50 statements with the highest prevalence in the HEEDB for MGH data, including their corresponding 12SL codes, human-readable descriptions, and counts. The statements encompass various elements, including diagnoses, rhythms, conjunctions, ECG lead names, and brackets and other symbols, all of which contribute to the formation of a human-readable statement generated by the 12SL software.

| 12SL Code | Statement | Count |
| --- | --- | --- |
| 1699 | Abnormal ECG | 7,361,517 |
| 22 | Normal sinus rhythm | 4,115,214 |
| 831 | , age undetermined | 2,170,034 |
| 1140 | Nonspecific T wave abnormality | 2,00,4701 |
| 19 | Sinus rhythm | 1,796,277 |
| 177 | With | 1,685,402 |
| 1684 | Normal ECG | 1,619,847 |
| 410 | Low voltage QRS | 1,330,445 |
| 244 | Fusion complexes | 1,288,987 |
| 390 | Indeterminate axis | 1,250,197 |
| 21 | Sinus bradycardia | 1,138,840 |
| 231 | Premature ventricular complexes | 1,130,305 |
| 1665 | ( | 1,129,759 |
| 1666 | ) | 1,129,759 |
| 411 | Pulmonary disease pattern | 927,790 |
| 23 | Sinus tachycardia | 902,408 |
| 372 | Left axis deviation | 851,802 |
| 1687 | Otherwise normal ECG | 826,410 |
| 780 | Inferior infarct | 798,145 |
| 161 | Atrial fibrillation | 767,742 |
| 299 | Undetermined rhythm | 732,518 |
| 542 | Minimal voltage criteria for LVH, may be normal variant | 705,711 |
| 1680 | Possible | 679,705 |
| 1141 | Nonspecific ST and T wave abnormality | 668,763 |
| 533 | Cornell product | 616,603 |
| 178 | Or | 601,429 |
| 530 | R in aVL | 591,281 |
| 101 | With 1st degree AV block | 575,095 |
| 1693 | Borderline ECG | 561,088 |
| 1100 | ST | 535,730 |
| 440 | Right bundle branch block | 514,185 |
| 740 | Anterior infarct | 474,416 |
| 1682 | Cannot rule out | 462,174 |
| 211 | With occasional | 432,974 |
| 380 | Rightward axis | 405,490 |
| 700 | Septal infarct | 393,446 |
| 1683 | , | 392,131 |
| 267 | Junctional rhythm | 387,313 |
| 1160 | T wave abnormality, consider lateral ischemia | 356,454 |
| 1146 | QTcB ≥ 480 ms | 355,159 |
| 1673 | *** Poor data quality, interpretation may be adversely affected | 350,927 |
| 251 | With sinus arrhythmia | 333,597 |
| 222 | Premature atrial complexes | 324,974 |
| 541 | Left ventricular hypertrophy | 321,334 |
| 482 | Nonspecific intraventricular block | 319,421 |
| 33 | Accelerated | 277,351 |
| 445 | Incomplete right bundle branch block | 266,564 |
| 360 | Left atrial enlargement | 262,051 |
| 171 | With rapid ventricular response | 252,983 |
| 1143 | Prolonged QT | 250,326 |

**Table 3.** This table lists the 50 statements with the highest prevalence in the HEEDB for Emory data, including their corresponding 12SL codes, human-readable descriptions and counts.

| 12SL Code | Statement | Count |
| --- | --- | --- |
| 1699 | Abnormal ECG | 1,297,906 |
| 831 | , age undetermined | 1,143,262 |
| 22 | Normal sinus rhythm | 682,788 |
| 1680 | Possible | 651,936 |
| 700 | Septal infarct | 580,815 |
| 760 | Lateral infarct | 425,489 |
| 820 | Anterolateral infarct | 230,557 |
| 520 | Right ventricular hypertrophy | 200,551 |
| 445 | Incomplete right bundle branch block | 191,208 |
| 21 | Sinus bradycardia | 189,975 |
| 1682 | Cannot rule out | 188,883 |
| 410 | Low voltage QRS | 174,791 |
| 19 | Sinus rhythm | 170,730 |
| 372 | Left axis deviation | 147,536 |
| 740 | Anterior infarct | 127,294 |
| 780 | Inferior infarct | 120,966 |
| 810 | Anteroseptal infarct | 120,699 |
| 23 | Sinus tachycardia | 108,889 |
| 1666 | ) | 105,221 |
| 1665 | ( | 105,221 |
| 530 | R in aVL | 97,627 |
| 101 | With 1st degree AV block | 91,620 |
| 161 | Atrial fibrillation | 88,573 |
| 231 | Premature ventricular complexes | 78,863 |
| 542 | Minimal voltage criteria for LVH, may be normal variant | 73,897 |
| 177 | With | 72,794 |
| 482 | Nonspecific intraventricular block | 65,658 |
| 440 | Right bundle branch block | 64,964 |
| 1160 | T wave abnormality, consider lateral ischemia | 62,600 |
| 1100 | ST | 57,750 |
| 211 | With occasional | 56,935 |
| 251 | With sinus arrhythmia | 54,769 |
| 1170 | T wave abnormality, consider inferior ischemia | 50,993 |
| 222 | Premature atrial complexes | 50,500 |
| 1340 | *** Critical test result: | 41,691 |
| 1143 | Prolonged QT | 40,725 |
| 1140 | Nonspecific T wave abnormality | 36,538 |
| 470 | Left anterior fascicular block | 35,114 |
| 1684 | Normal ECG | 31,679 |
| 1150 | T wave abnormality, consider anterior ischemia | 29,813 |
| 541 | Left ventricular hypertrophy | 29,242 |
| 171 | With rapid ventricular response | 28,167 |
| 212 | With frequent | 23,383 |
| 181 | With premature ventricular or aberrantly conducted complexes | 21,063 |
| 1683 | , | 20,472 |
| 544 | With repolarization abnormality | 19,683 |
| 221 | Premature supraventricular complexes | 18,448 |
| 162 | Atrial flutter | 18,178 |

The statement numbers include various components such as ECG diagnoses, morphology, rhythms, lead names, text symbols (e.g., brackets, delimiters), measurements (e.g., “QTcB *≥* 480 ms”), conjunctions (e.g., “with,” “or,” “and”), and descriptive words like “present,” “accelerated,” and “blocked.” Together, these elements comprise the 12SL language model to form human-readable diagnosis reports. For example, the following statement consists of the codes: 19, 177, 231, 178, 244, 390, 411, 700, 831, 1100, 1150, and 169, which map to the diagnosis: Sinus rhythm, with, premature ventricular complexes, or, fusion complexes, Indeterminate axis, Pulmonary disease pattern, Septal infarct,, age undetermined, ST & T wave abnormality, consider anterior ischemia, Abnormal ECG. Annotations are available for 10,471,531 ECG recordings, leaving 136,886 unlabeled recordings from MGH. For the EUH dataset, 1,268,277 recordings are annotated, with 184,687 remaining unlabeled.

## Technical Validation

The ECG records were obtained in a clinical environment, during routine care guided by specialized personnel using the MUSE ECG system facilitating acquisition, storage, and review of ECG data, enabling consistent recordings across the dataset. The HEEDB data was checked for completeness and includes the non-preprocessed raw ECG waveforms in WFDB format coupled with demographics and 12SL statements. Data was de-identified following the Safe Harbor method. ECG recordings contain various types of standard ECG artifacts such as missing leads, missing parts of signals, or noise.

## Usage Notes

The HEEDB dataset can be downloaded from the Brain Data Science Platform (BDSP) by following the instructions provided at Harvard-Emory ECG database website^4,5^ (https://bdsp.io/content/heedb/2.0/). The data are made available under an Attribution - NonCommercial - NoDerivatives (CC BY-NC-ND) license. This license enables reusers to copy and distribute the material in any medium or format in **unadapted** form only, for **noncommercial purposes** only, and only so long as **attribution is given to the creator**.

## Data Availability

All data produced are available online with credentialed access at https://bdsp.io/content/heedb/1.0/ and shortly will be available publicly.

## Code availability

The downloaded waveform data files can be accessed using WFDB library functions, applications, and toolboxes in Python^6^ and MATLAB^7^ or can be directly loaded into MATLAB. No custom code was developed for this study.

## Acknowledgements

Publication of HEEDB is supported by a grant (R01HL161253) from the National Heart Lung and Blood Institute (NHLBI) of the NIH to Massachusetts General Hospital, Emory University, Stanford University, Kaiser Permanente, Boston Children’s Hospital, and Beth Israel Deaconess Medical Center. Publication of the HEEDB is also supported by the National Institute of Biomedical Imaging and Bioengineering (NIBIB) under NIH grant number R01EB030362, and an unrestricted funds from Emory University. Q.L., M.R. and G.D.C. are supported in part by unrestricted funding from AliveCor Inc.

## Author contributions statement

Z.K. provided technical validation of the database, statistical analysis of the database, processing of the annotations and drafted the manuscript; Q.L. contributed to statistical analysis of the data; C.R. contributed to data collection; J.M.V contributed to data collection, developed infrastructure for data storage; M.G. contributed to data collection; A.G. conducted preliminary analyses of the data; J.R. conducted preliminary analyses of the data; S.H. conducted preliminary analyses of the data; A.A. contributed to data collection; D.E.A. provided insights into the clinical relevance of the data; J.X secured data annotations; A.P.secured data annotations; R.S. contributed to the design and implementation of the data collection processes as well as overseeing the technical validation of the database and contributed to the writing; M.A.R. contributed to the design and implementation of the data collection processes, contributed to the writing and led; M.B.W. secured the funding, designed the database, contributed to the design and implementation of the data collection processes, and contributed to the writing; G.D.C. secured the funding, designed the database, provided intellectual input on processing and labeling, contributed to the manuscript and led the project. All authors read the manuscript.

## Competing interests

M.B.W. is a co-founder, scientific advisor, and consultant to Beacon Biosignals and has a personal equity interest in the company. D.E.A. is the founder and Chief Medical Officer of AliveCor Inc. and holds personal equity in the company. J.X. is a research fellow at Alivecor Inc. and also holds personal equity in the company. G.D.C. holds significant stock in AliveCor Inc. The other authors report no competing interests.

## Notes

### Author Declarations

Ethics committee of Emory University gave ethical approval for this work.

### Summary of Updates

We updated the license to: CC BY-NC-ND

